# NeuroXNet: Creating A Novel Deep Learning Architecture that Diagnoses Neurological Disorders, Finds New Blood Biomarkers, and Assesses Surgical, Drugs, and Radiation Treatment Plans Using Medical Imaging and Genomic Data

**DOI:** 10.1101/2021.12.13.21267728

**Authors:** Vaibhav Mishra

## Abstract

Neurodegenerative diseases and cancerous brain tumors cause millions of patients worldwide to be fatally ill and face cognitive impairment each year. Current diagnosis and treatment of these neurological conditions take many days, are sometimes inaccurate, and use invasive approaches that could endanger the patient’s life. Thus, this study’s purpose is the creation of a novel deep learning model called NeuroXNet, which uses MRI images and genomic data to diagnose both neurodegenerative diseases like Alzheimer’s disease, Parkinson’s disease, and Mild Cognitive Impairment as well as cancerous brain tumors, including glioma, meningioma, and pituitary tumors. Moreover, the model helps find novel blood biomarkers of differentially expressed genes to aid in diagnosing the six neurological conditions. Furthermore, the model uses patient genomic data to give additional recommendations for treatment plans that include various treatment approaches, including surgical, radiation, and drugs for higher patient survival for each class of the disease. The NeuroXNet model achieves a training accuracy of 99.70%, a validation accuracy of 100%, and a test accuracy of 94.71% in multi-class classification of the six diseases and normal patients. Thus, NeuroXNet reduces the chances of misdiagnosis, helps give the best treatment options, and does so in a time/cost-efficient manner. Moreover, NeuroXNet efficiently diagnoses diseases and recommends treatment plans based on patient data using relatively few parameters causing it to be more cost and time-efficient in providing non-invasive approaches to diagnosis and treatment for neurological disorders than current procedures.

## I. Introduction

### 1.1 Background

Neurological disorders continue to affect millions of people worldwide, with diseases leading to loss of cognitive function, a decline in memory, and even death. These diseases contribute to nearly a trillion dollars of healthcare spending and drastically change the lives of those affected. With the advent of new medical imaging and computational techniques, it has become possible to use large amounts of imaging data to build and train deep learning models that can diagnose many diseases with high accuracy rates using clinical tests and medical imaging tests like MRI. Some of the most common neurodegenerative disorders include Alzheimer’s disease, Parkinson’s disease, and Mild Cognitive Impairment. In the United States, approximately 6.2 million people are affected by AD [6], 1 million people are affected with PD [7], and 11 million people are affected with MCI [8].

Alzheimer’s Disease (AD) is the most common neurodegenerative disease that starts with MCI like symptoms of memory loss and cognitive decline and later evolves to damage neurons responsible for carrying out essential functions affecting everyday life. Eventually, AD can result in the death of a patient. Some possible causes of AD include beta-amyloid plaques that form outside the neurons and tau tangles inside neurons, which have been observed inside several AD patients [6]. MRI images can diagnose AD earlier by locating critical parts of the brain that signal a higher possibility of AD. Some of the primary areas used to diagnose AD in the brain are the hippocampus and the parietal lobes. However, in patients with AD, the MRI will clearly show a decrease in the size of the hippocampus signaling a higher probability of AD. The parietal lobe is also a signal that radiologists look for in diagnosing AD through MRIs. The parietal lobe is responsible for sensory information and visual processing in our bodies. A size reduction in a patient’s parietal lobe also signals a chance of AD in a patient. [18] Therefore, MRIs must be adequately interpreted and used to give an early diagnosis of patients to prevent AD from progressing. Mild Cognitive Impairment (MCI) is a neurological condition with symptoms that show loss of certain cognitive functions like memory or the ability to speak. MCI can convert to AD or other types of dementia over a few years, remain stabilized as MCI, or revert to a normal functioning brain. The most common treatment of MCI is Aducanumab, a drug used to treat AD. [8] One major problem of diagnosis of MCI is that MCI can be similar in symptoms to normal cognitive decline due to old age in many people, which causes many patients to ignore the signs and leave MCI untreated. This is harmful as MCI can later develop into AD over time. Therefore, it is crucial to diagnose and treat MCI in its early stages to avoid more cognitive loss in patients.

Parkinson’s Disease (PD) is a neurological disease that primarily damages the motor neurons disturbing a patient’s movement ability. PD usually results in patients having worse conditions over more extended periods. However, PD can be treated with certain medications like Carbidopa-levodopa [10]. MRIs can help detect specific abnormalities to brain structures that signal PD in PD patients. Particularly in PD, there are significant changes seen in the right parietal cortex, the putamen, and brain structures responsible for motor actions like the mesencephalic locomotor region. The right parietal lobe is chiefly responsible for various functions of the body, including taste, touch, and the somatic sensory cortex within it. The putamen has the function of controlling movement, and as such, these areas are critical for PD diagnosis [19].

Glioma tumors are located in the central nervous system, and various glial cells are found throughout the peripheral nervous system, including astrocytes, oligodendrocytes, and microglia. Glioma tumors include glioblastoma multiforme, astrocytoma, and oligodendroglioma. Oligodendroglioma can occur in either the temporal or frontal lobes and cause seizures and motor inability. This study focuses on more lethal kinds of brain tumors, including anaplastic astrocytoma (which appears more commonly in the cerebral hemisphere) and glioblastoma multiforme (which starts in the cerebrum in astrocyte cells) [26]. Pituitary tumors primarily occur in the anterior portions of the pituitary gland. Pituitary tumors cause vertigo, fatigue, and problems with visual perception [26]. Pituitary tumors are generally treated through surgery using transsphenoidal resection [26]. Meningioma tumors occur on the meninges of the brain and can cause vision problems as well as seizures [26].

### 1.2 Previous Literature

Through the usefulness of large amounts of data and advancements in medical imaging research, it has become possible to diagnose and even treat patients with diseases in various fields of medicine. A large amount of medical imaging research using MRI data has been used to classify AD from normal patients. For instance, Al-Khuzaie et al. [4] developed a new model, AlzNet, which achieved an accuracy of 99.30% in diagnosing AD from normal using 2D MRI slices. One of the highest accuracies was achieved in AD diagnosis using machine learning models. Another paper [11] trained a CNN model to classify AD, normal, and MCI patients using 3D MRI with an accuracy of 52.3%.

Medical imaging research has also been extended into PD classification from normal patients. Multiple models have been developed that incorporate a machine learning approach to classify PD from MRI imaging. A research team led by Long [1] developed a model in which accuracy of 86.96% was achieved in classifying and diagnosing PD patients from normal patients. In [56], computational tools identify novel blood biomarkers in PD patients using gene ontology analysis and protein interactions.

However, relatively few machine learning models have performed multiclass diagnosis in neurological disorders. A multiclass approach to classifying multiple neurodegenerative diseases was studied in very few papers, notably in [14], where AD, PD, and CN were classified with an accuracy of 90% for AD, 90% for control from ADNI, 89% for control from PPMI, and 90% for PD using transfer learning on the VGG19 model which performed the best out of the ResNet 50, Inception Net, and VGG16 model which were also tested in the paper and in another article by Tong et al. [15], where a five-class model was proposed that achieved a 75.2% that classified AD and other dementia-like diseases using the RUSBoost algorithm. Even in these studies, the model was limited to classifying only neurodegenerative diseases in lesser than five classes. Therefore, this study aims to solve the problem of multiclass diagnosis and treatment of neurological disorders.

### 1.3 Purpose of Study

This study proposes a novel deep learning architecture, NeuroXNet, which performs multiclass diagnosis of AD, PD, MCI, glioma, meningioma, pituitary, and normal patients. In addition, NeuroXNet generates recommendations for treatment based on disease classification from its convolutional neural network (CNN) model combined with the patient’s genomic and clinical data. These recommendations include treatment plans for surgery, radiation, or drug therapy. This is the first CNN model which creates a novel architecture to study multiclass diagnosis instead of relying on previously built models like ResNet50 or VGG16. Furthermore, this model discovers new blood biomarkers using genomic data to further aid in diagnosing neurological conditions and reducing the chance of misdiagnosis. These genes can also be further studied to find possible targets for treatment research. Moreover, this model is the first that combines diagnosis with treatment plans for neurological disorders. Therefore, this model has great potential to be used in neurological medicine and provide a low-cost, efficient, and quick solution to patients worldwide.

## II. Materials and Methods

### 2.1 Data Acquisition and Description for MRI Classifier

Data of AD and MCI patients used in the paper was obtained from the Alzheimer’s Disease Neuroimaging Initiative (ADNI), a free open source database (adni.loni.usc.edu), and data for the PD and normal patients was obtained from the Parkinson’s Progression Markers Initiative (PPMI) database (ppmi-info.org). Data for MRI of patients with glioma, meningioma, and pituitary tumors were acquired from Kaggle’s Brain Tumor Dataset [22]. For each of the seven AD, PD, MCI, Pituitary Tumor, Glioma Tumor, Meningioma Tumors, and Normal Patients, 1000 Brain MRI images were collected NIFTI format from the respective databases for a total of 7000 images. All the MRIs were T1-weighted and equally distributed to reduce model bias. These images were then converted to png format using the MRIcro tool. The images were split into training/validation/test folders using a 70%/30%/10% ratio. Table 1 shows the number of patient MRIs that were split into Training, Validation, and Test sets for each disease class.

**Table 1:** Number of Patients per Data Set Folder.

| Table 1: Data Split Into Training, Validation, and Test Sets |  |  |  |  |
| --- | --- | --- | --- | --- |
|  | Training Set | Validation Set | Test Set | Total |
| Alzheimer's Disease | 700 | 300 | 100 | 1000 |
| Parkinson's Disease | 700 | 300 | 100 | 1000 |
| Mild Cognitive Impairment | 700 | 300 | 100 | 1000 |
| Glioma Tumor | 700 | 300 | 100 | 1000 |
| Meningioma Tumor | 700 | 300 | 100 | 1000 |
| Pituitary Tumor | 700 | 300 | 100 | 1000 |
| Normal Control | 700 | 300 | 100 | 1000 |
| Total | 4900 | 2100 | 700 | 7000 |

The demographic data including age and gender for the neurodegenerative classes is shown below in Table 2:

**Table 2:**
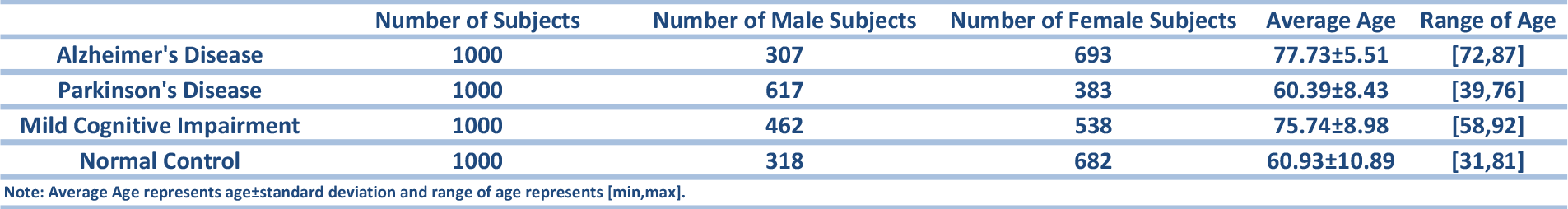
Demographic Data for Neurodegenerative Diseases.

**Table 3:** Disease Classification Report.

| Table 3: Classification Report for Diseases |  |  |  |  |
| --- | --- | --- | --- | --- |
|  | Precision | Recall | F1-score | Support |
| AD | 0.92 | 0.92 | 0.92 | 100 |
| MCI | 0.97 | 0.97 | 0.97 | 100 |
| PD | 0.92 | 0.92 | 0.92 | 100 |
| Glioma | 0.97 | 0.97 | 0.97 | 100 |
| Meningioma | 0.96 | 0.96 | 0.96 | 100 |
| Pituitary | 0.95 | 0.95 | 0.95 | 100 |
| Normal | 0.94 | 0.94 | 0.94 | 100 |

**Table 4:** Model Classification Report.

| Table 4: Classification Report for Whole Model |  |  |  |  |
| --- | --- | --- | --- | --- |
|  | Precision | Recall | F1-Score | Support |
| Accuracy |  |  | 0.95 | 700 |
| Macro Average | 0.95 | 0.95 | 0.95 | 700 |
| Weighted Average | 0.95 | 0.95 | 0.95 | 700 |

**Table 5:** Surgical Treatment Summary for NeuroXNet.

| Table 5: Surgical Treatment Summary for NeuroXNet |  |  |
| --- | --- | --- |
|  | Determining Factor | Result |
| PD | Early Stages of PD | Deep Brain Stimulation |
|  | Late Stages of PD | Duopa |
| MCI | Patient Needs Pain Control | Transcutaneous Electrical Nerval Stimulation (TENS) |
|  | Does Not Need Pain Control | Use Medicine or Alternative Approaches |
| Pituitary | Tumor grater than or equal to 10mm in diameter size | Craniotomy |
|  | Tumor less than 10mm in diameter size | Endoscopic Transnasal Transsphenoidal Approach |
| Meningioma | Tumor found in ventricular chambers | Neuroendoscopic Surgery |
|  | Tumor grater than or equal to 10mm in diameter size | Craniotomy |
|  | Tumor less than 10mm in diameter size | Endoscopic Endonasal |
| Glioma | Tumor has caused build up of CSF | Shunt Placement |
|  | Surgery Needed to Test Tumor Removal Effects | Brain Mapping and Awake Surgery |
|  | Surgery Needed to Test Tumor Tissue | Surgical Biopsy |
|  | Surgery Required to Remove Tumor Without Any Functionality Tests | Surgical Resection |
| AD | Patient Needs Pain Control | Transcutaneous Electrical Nerval Stimulation (TENS) |
|  | Patient in Early Stages of AD | Cerebrospinal Fluid Shunting |
|  | Patient in Moderate Stage of AD | Gene Therapy |
|  | Patient in Late Stage of AD | Intraventricular Infusion |

**Table 6:**
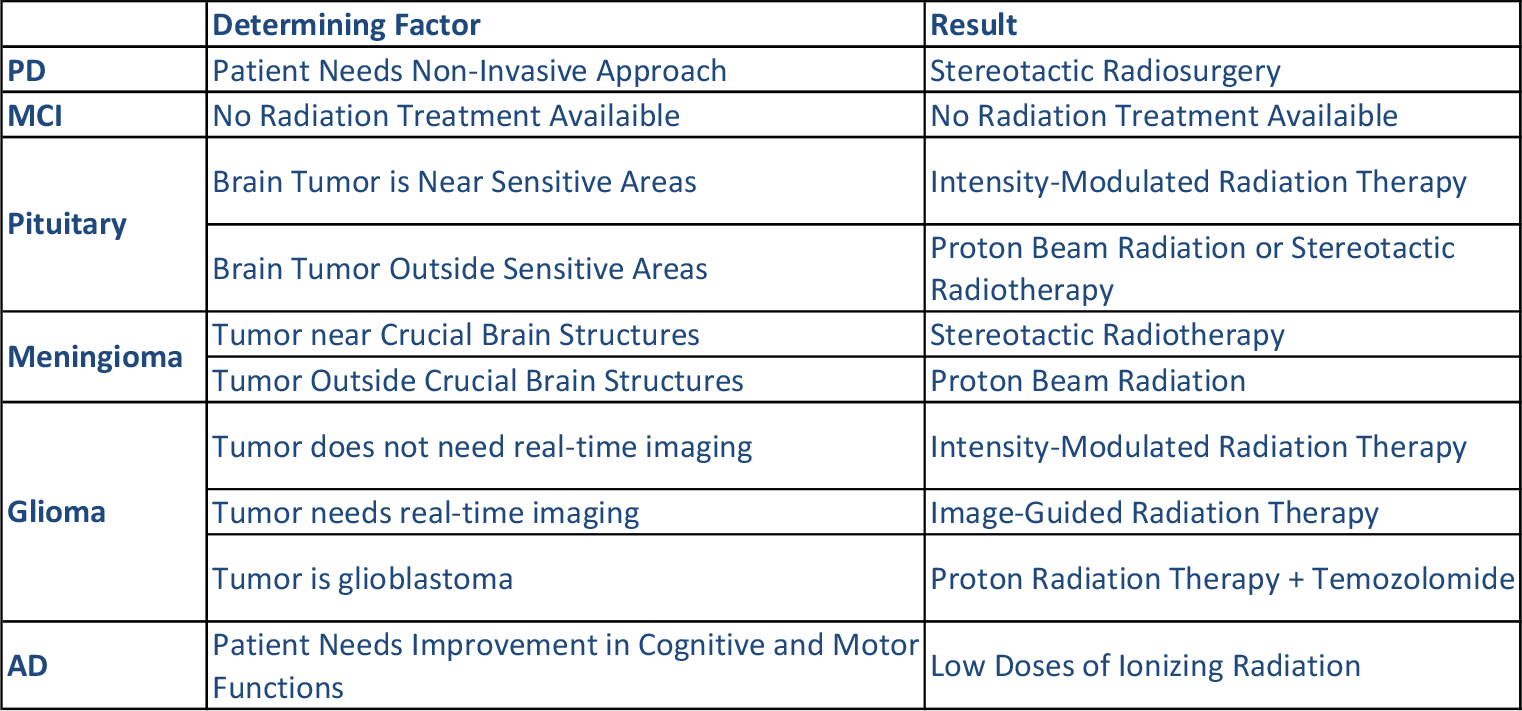
Radiation Treatment Summary for NeuroXNet.

**Table 7:** Drug Treatment Summary for NeuroXNet.

| Table 7: Drug Treatment Summary for NeuroXNet |  |  |
| --- | --- | --- |
|  | Determining Factor | Result |
| PD | Patient Does Not Have DNA Methylation Occuring In Brain Cells | Levodopa+Carbidopa |
|  | Patient Has DNA Methylation Occuring In Brain Cells | Levodopa+Carbidopa+Drugs used to prevent Levodopa-Induced Dyskenesia |
| MCI | No Medication Treatment Available | No Medication Treatment Available |
| Pituitary | Pituitary tumor secretes excess cortisol | Ketoconazole |
|  | Pituitary tumor secretes excess growth hormone | Pegvisomant |
|  | Pituitary tumor secretes excess prolactin | Bromocriptine+Cabergoline |
| Meningioma | Patient be subjected to surgery | Use surgical options |
|  | Patient cannot be subjected to surgery | Interferon or Bevacizumab or Hydroxyurea |
| Glioma | Patient has over expression of MGMT gene | Temozolomide+Radiation or Temozolomide+MGMT-gamma-delta modified Tcells |
|  | Patient does not have over expression of MGMT gene | Temozolomide |
| AD | Patient needs overall improvement in all AD symptoms | Aducanumab |
|  | Patient needs overall improvements only in memory and thinking functions | Donepezil or Rivastigmine |
|  | Patient needs overall improvements in cognitive functions | Memantine |
|  | Patient needs overall improvement in behavioral functions | Orexin receptor antagonists with suvorexant |

### 2.2 Data Preprocessing for Diagnosis

After being split into their respective folder, each MRI image was normalized by rescaling the size of each image. Then, data augmentation was applied, including shearing, zooming, and flipping the data to decrease overfitting and improve the model’s overall accuracy. The data was preprocessed to reduce the amount of data bias the model gains and helps make the model regularize to fit all ranges of MRI images.

### 2.3 Deep Learning Models Overview

Convolutional Neural Networks (CNNs) are a class of deep neural networks part of supervised machine learning that are most widely used for visual image processing, including medical imaging. CNN’s use multiple layers to take input images, find patterns within them, and learn from certain features within the pictures. They then use the learned weights and biases to output the image’s class. A convolutional layer is the centerpiece of the NeuroXNet model, which uses a set of weights and biases (also called factors) that learns over time.

### 2.4 Identification of Novel Biomarkers

Identifying biomarkers from blood, tissue, or another type of body cell for early detection of neurological diseases was one of the main focuses of this study. Blood biomarkers are analyzed with microarrays to gather data on gene expression for thousands of genes and subsequently used as a non-invasive neurological test for diagnosis and research in finding new treatments. This study utilized genomic datasets from Gene Expression Omnibus. The data used in this study consisted of the series GSE74385 [46, 47] for meningioma tumor classification, GSE31095 [48] for glioma tumors, GSE4488 [49] for pituitary tumors, GSE63063 [50] for MCI classification, GSE6613 [51, 52] for PD classification, and GSE4226 [53, 54] for AD classification. Each gene expression study was used to find differentially expressed genes that would serve as blood biomarkers for diagnosing the specific neurological disorder. The NeuroXNet model used to classify patients by genomic data.

Using GEO2R, the patients were split into two groups for each series number, with one group consisting of the specific disease class and the other group with normal patients. Then, all differentially expressed genes with a p-value of less than 0.05 were used to generate a protein interaction network using the STRING tool. The protein interaction network was then used to identify hub genes with a high number of nodes using the Cytoscape tool. The hub genes are the differentially expressed genes that are over-expressed in patients with certain neurological conditions and are potential biomarkers for the disease. These hub genes were sorted by degree (number of nodes connected to) and used for gene ontology analysis using the PANTHER tool, which gave the false positive rate, fold enrichment, and p-values for the specific genes present in biological, molecular, and cellular processes.

The fold change represents the ratio of the average gene expression in the experimental group vs. the control group. This study focused on the over-expressed genes with fold change values greater than 1.

### 2.5 NeuroXNet Model Description

NeuroXNet is the novel CNN model created in this study that uses patient MRI data and genomic data to diagnose and give treatment recommendations to patients, including surgical, radiation, and drug treatments to improve patient survivability. First, the model inputs MRI images, preprocesses them and sends them to the NeuroXNet Model. The model uses pre-trained weights to classify the patient’s MRI as AD, PD, MCI, glioma tumor, meningioma tumor, pituitary tumor, or normal. Next, genomic data of the patient is entered in as a secondary input and used to diagnose the patient again to ensure an accurate diagnosis. The model diagnoses patients using its pre-trained algorithm from blood biomarkers which it found beforehand. The classification information is then passed through the treatment tool of the NeuroXNet model, which matches the classified disease to its best treatment plan taking into account the susceptibility of the disease to drug treatments, radiation treatments, or surgical treatments starting from non-invasive options first that provide the most minor side effects to the patient and eliminating options that have low rates of survival based on patient’s genomic data. The process is shown in Figure 1 below:

**Figure 1:**
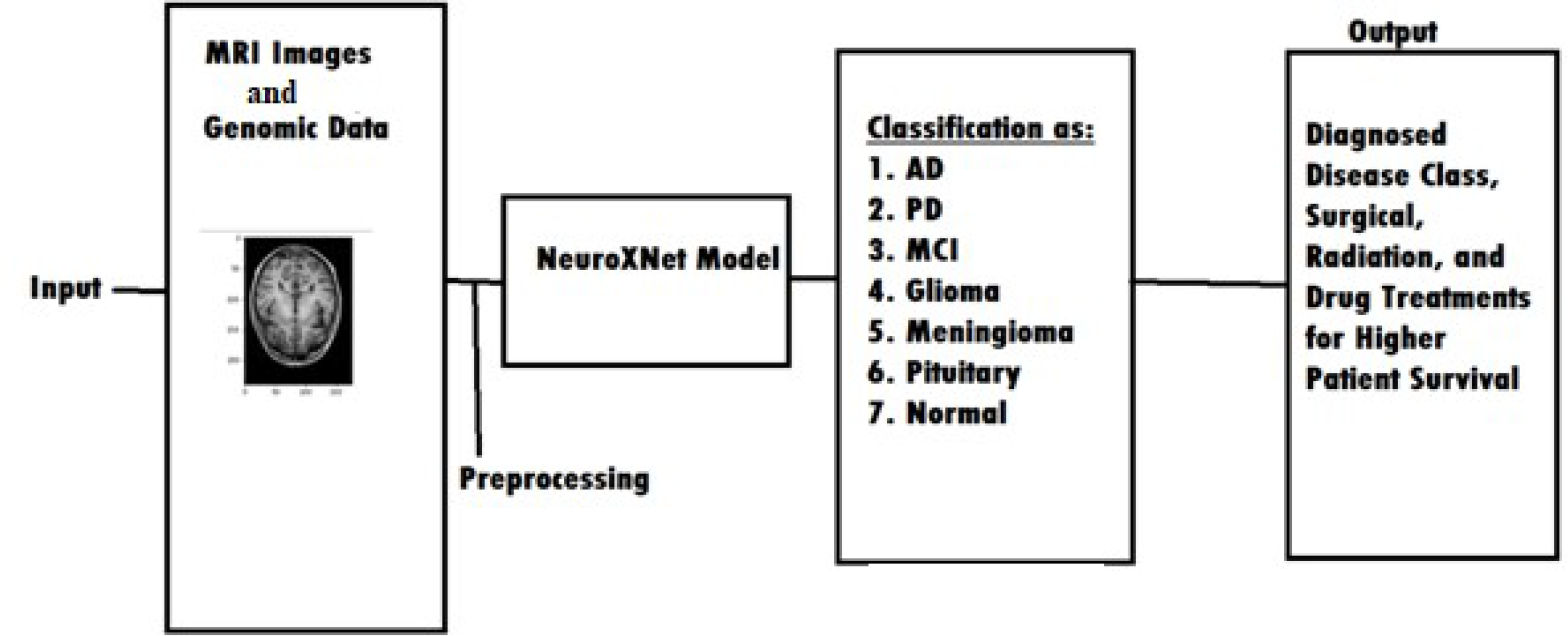
NeruoXNet Model Workflow Analysis.

**Figure 2:**
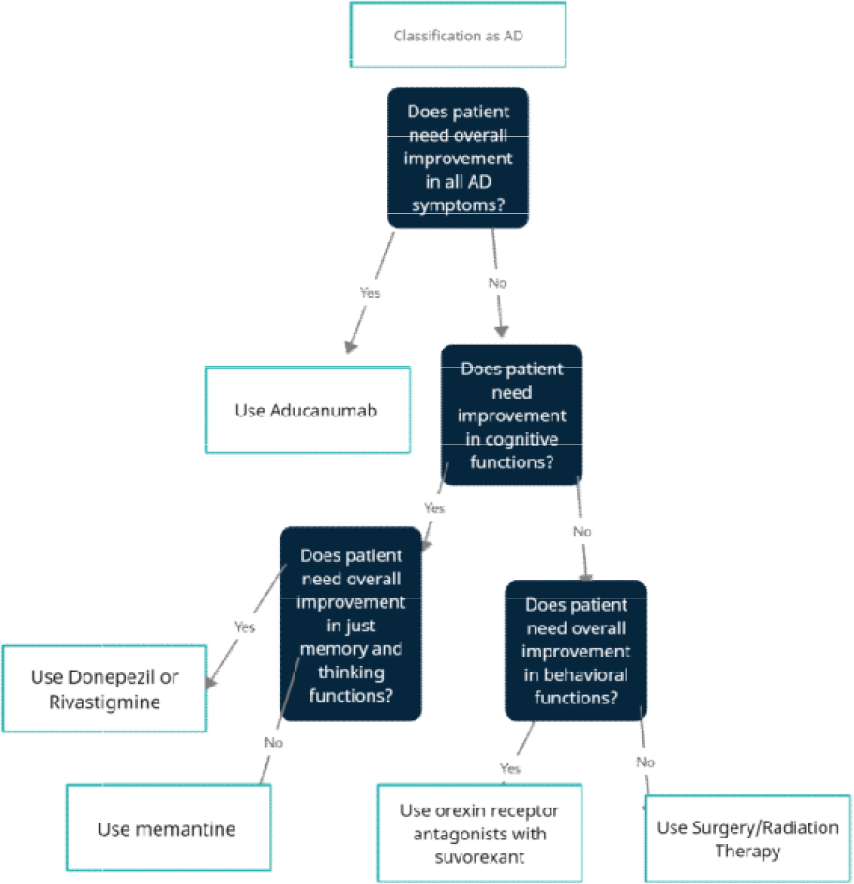
Example of NeuroXNet Algorithm for AD Drug Treatment.

**Figure 3:**
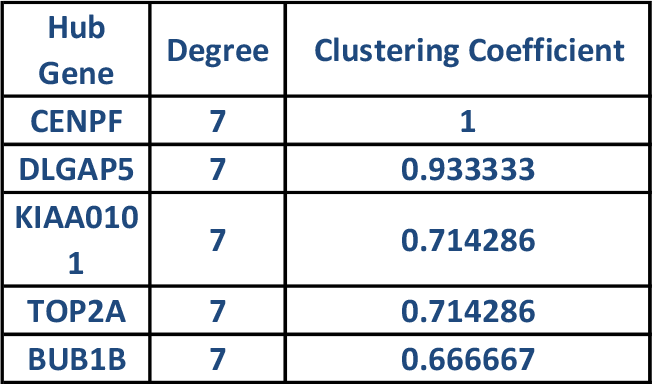
Hub Genes for PD.

**Figure 4:**
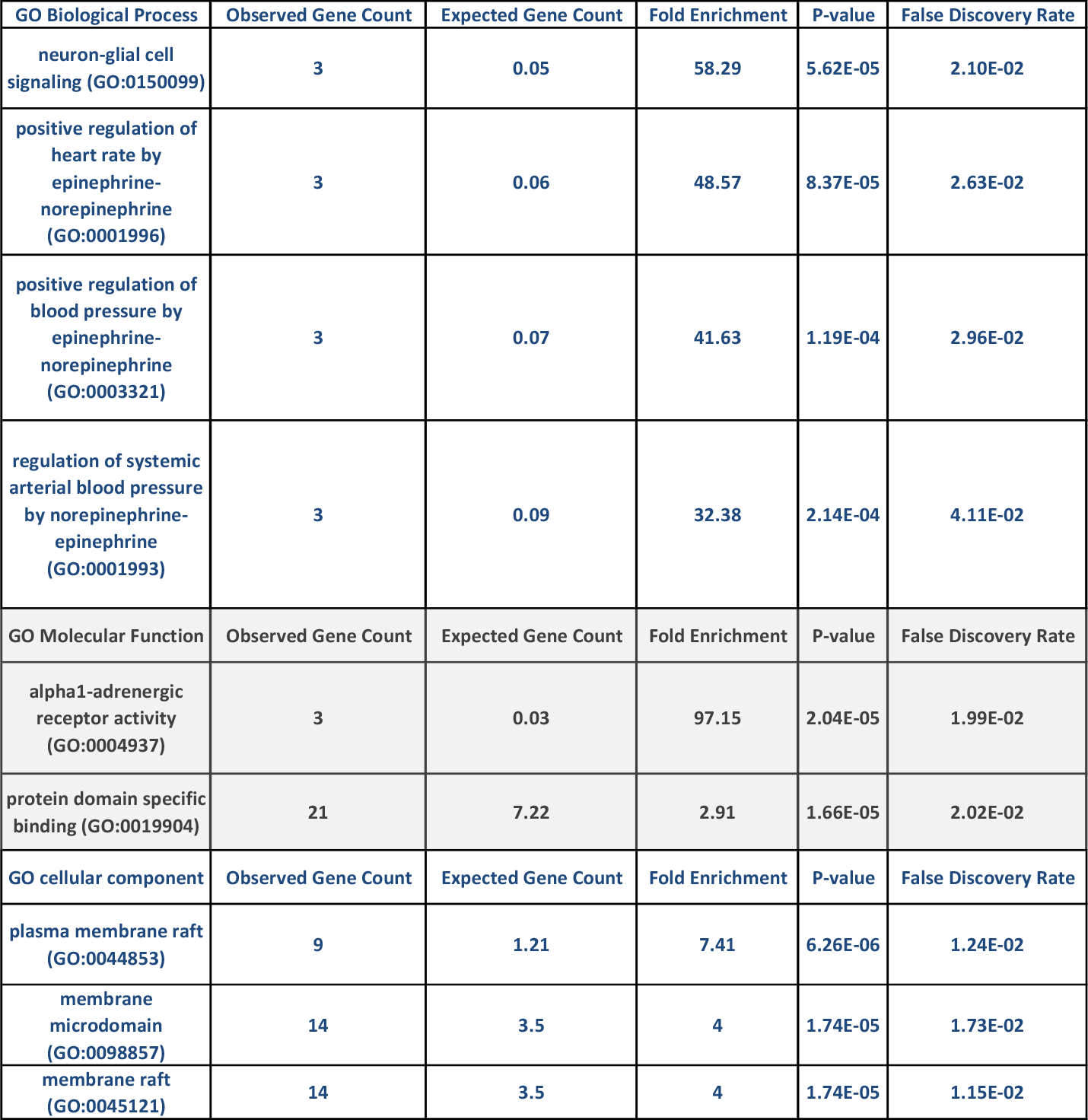
Biological, Molecular, and Cellular Functions from PD Over expressed Gene.

**Figure 5:**
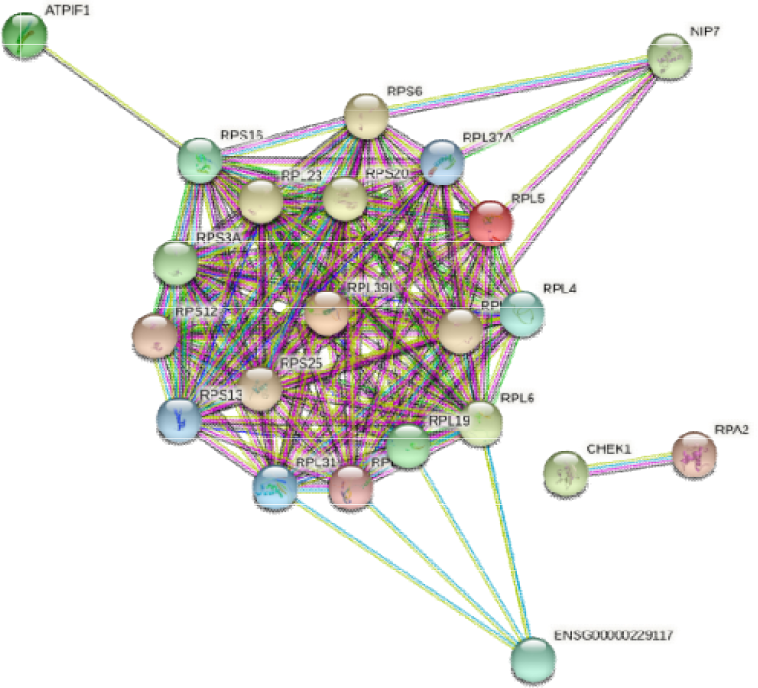
Protein Interaction Network of Biomarkers for AD.

**Figure 6:**
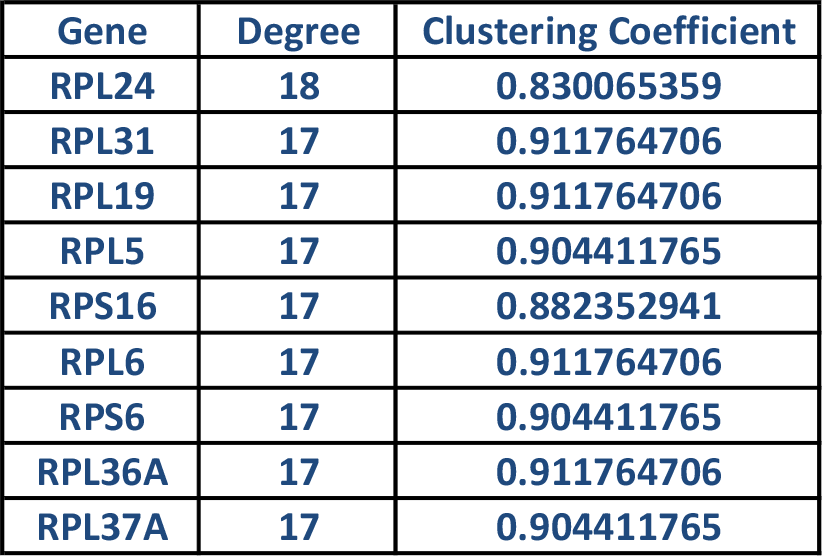
Hub Genes for AD.

**Figure 7:**
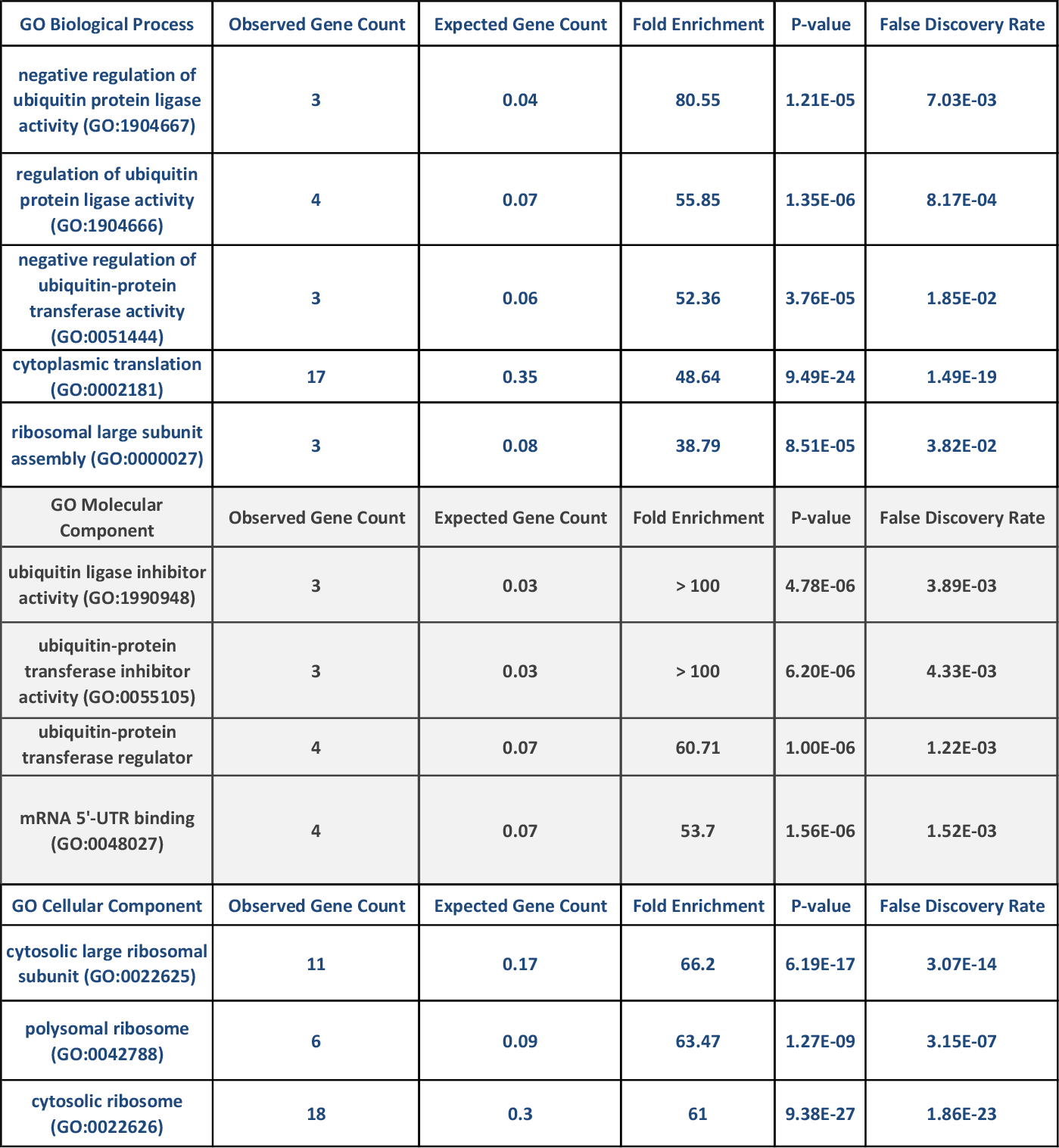
Biological, Molecular, and Cellular Functions from AD Over expressed Genes.

**Figure 8:**
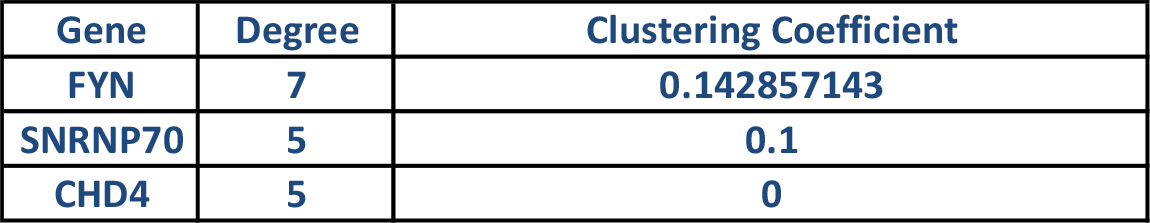
Hub Genes for MCI.

**Figure 9:**
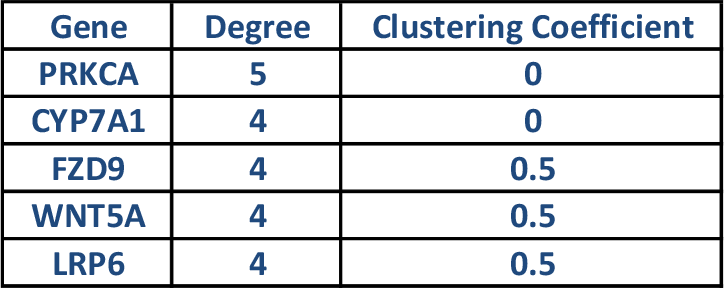
Hub Genes for Glioma Tumors.

**Figure 10:**
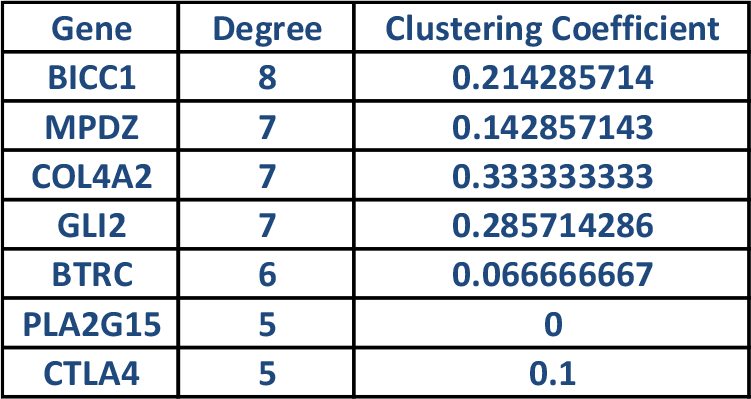
Pituitary Hub Genes.

**Figure 11:**
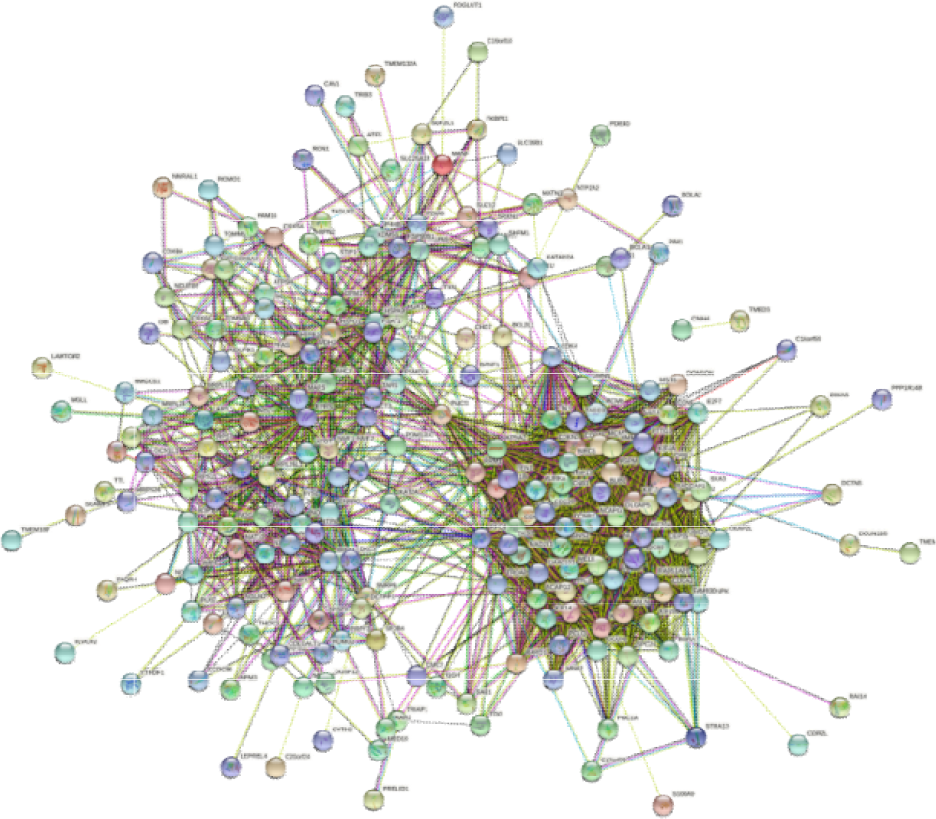
Protein Interaction Network for Meningioma Over Expressed Genes.

**Figure 12:**
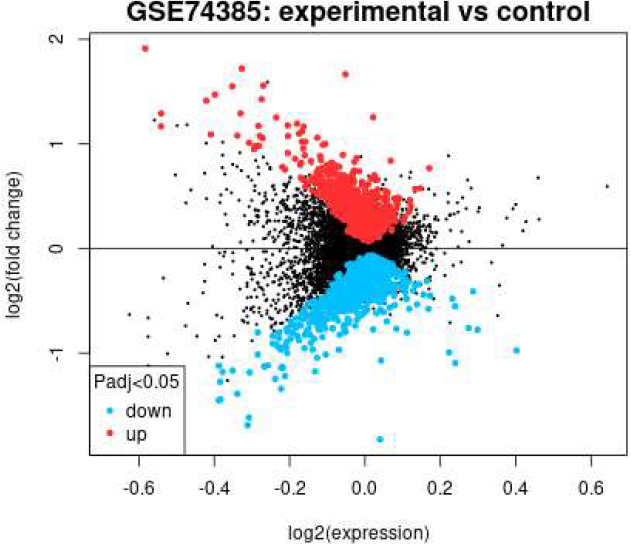
Mean Difference Plot for Meningioma Related Genes.

**Figure 13:**
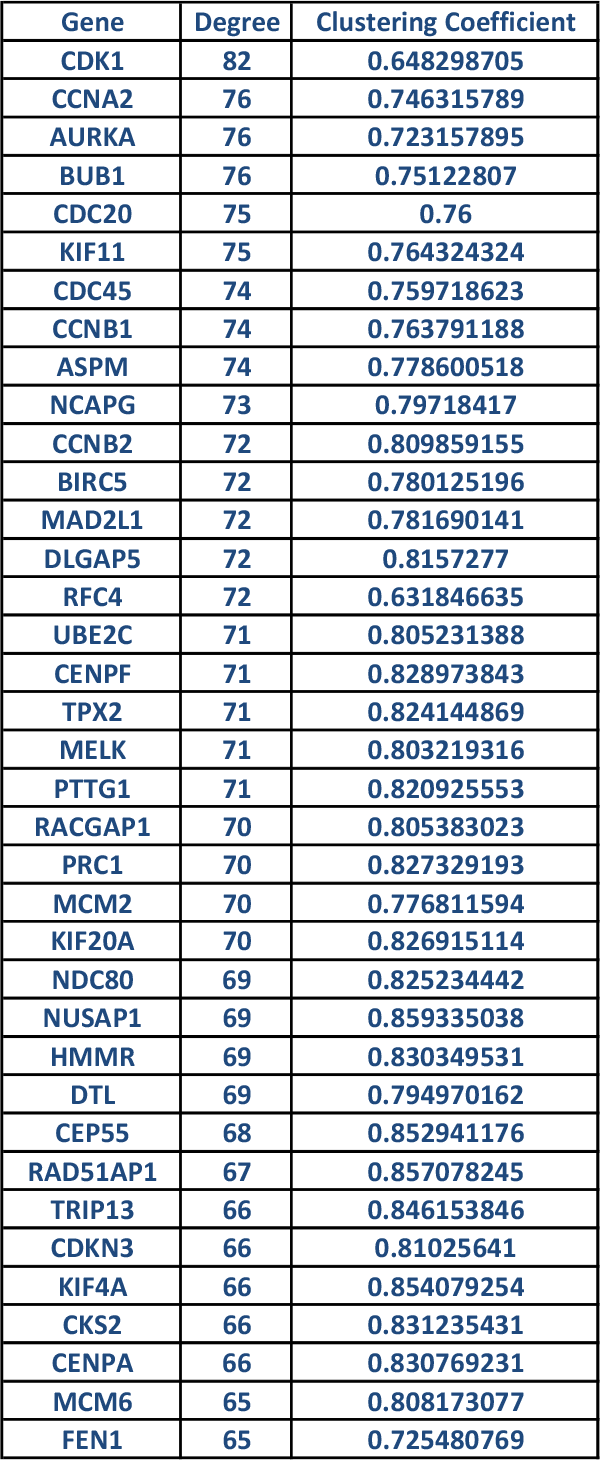
Meningioma Hub Genes.

**Figure 14:**
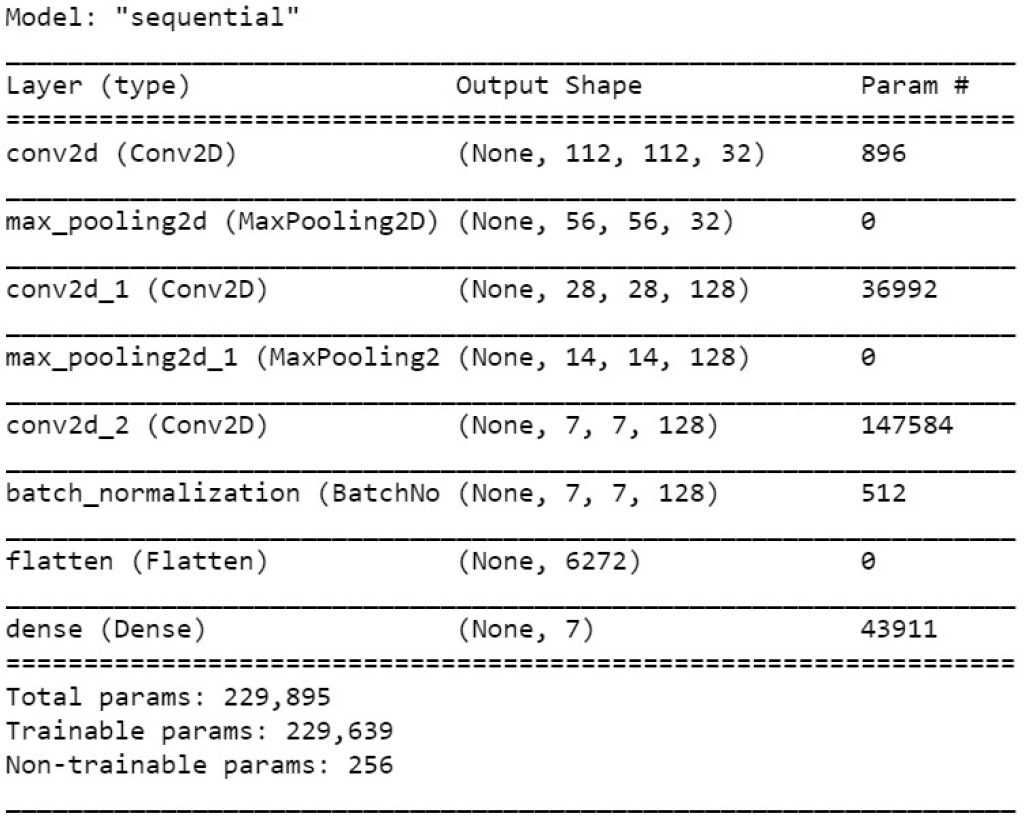
NeuroXNet Model Architecture for Disease Diagnosis.

**Figure 15:**
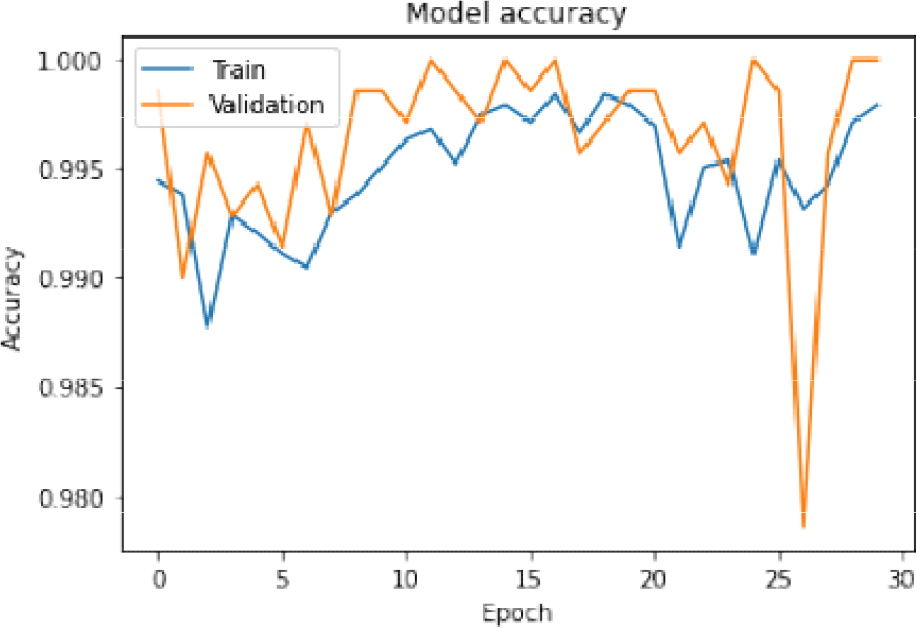
Training and Validation Model Accuracy.

**Figure 16:**
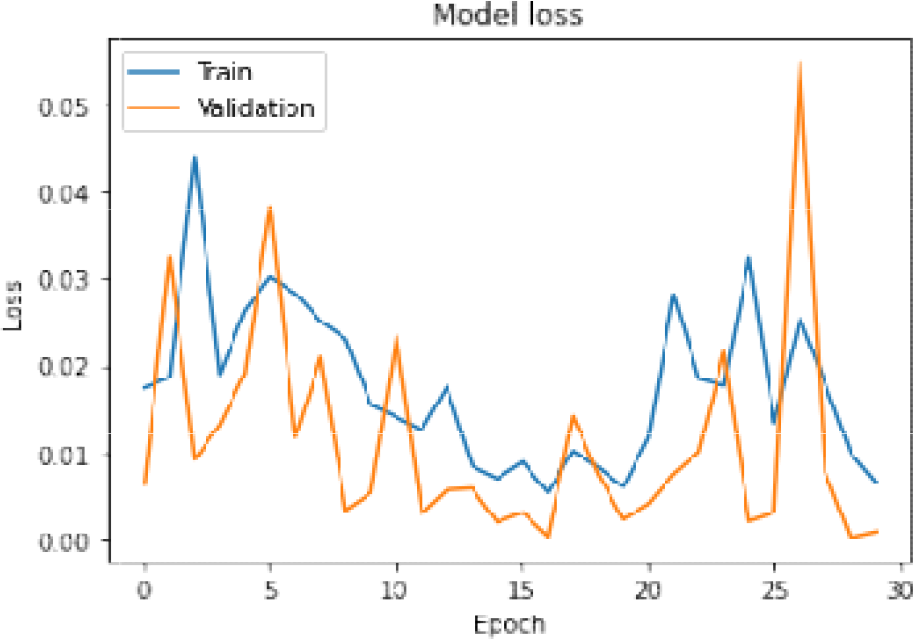
Training and Validation Model Loss.

**Figure 17:**
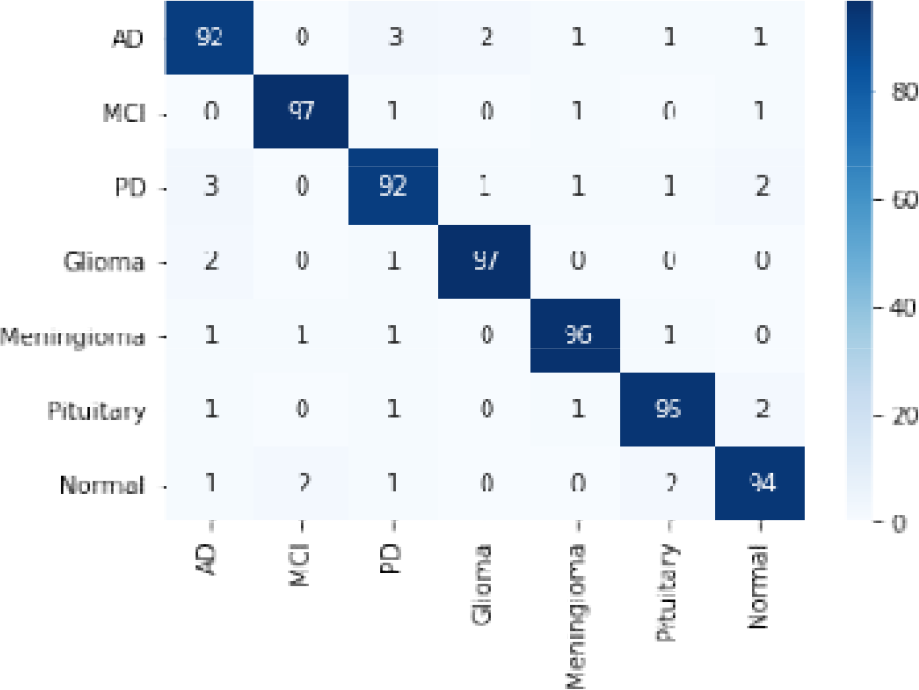
Model Confusion Matrix and Heatmap.

The next three sections detail the kind of algorithmic process that the NeuroXNet model goes through in suggesting treatment procedures for patients.

### 2.6 Surgical Treatment Procedures

For surgical procedures, NeuroXNet focuses on only widely used methods and can cause significant improvements to patient health. The model makes recommendations based on the diagnosed disease input from the result of its CNN model.

For patients diagnosed with PD, the model considers only the two main surgical methods used by surgeons: Deep Brain Stimulation (DBS) and Duopa. DBS is a surgical procedure that involves placing electrodes into the brain with an impulse generator battery that is connected to a wire that passes through the head and shoulder. DBS delivers electrical stimulation to the subthalamic nucleus and the globus pallidus internus, which control movements that cause tremors to reduce PD symptoms [28]. DBS is used for patients that are in the early stages of PD and experience uncontrolled tremors [28].

For patients that are diagnosed with MCI, the NeuroXNet model consists of mainly one surgical treatment procedure: Transcutaneous Electrical Nerval Stimulation (TENS). TENS is a medical procedure that involves applying an electrical current to electrons located on the skin in order to reduce pain in patients. TENS improves cognitive functioning, as well as counters, sleep disorders, and neural degeneration [30].

For patients that have pituitary tumors, the model either suggests a craniotomy or an endoscopic transnasal transsphenoidal approach [31]. A craniotomy is performed by making an incision on the scalp, after which a portion of the skull is removed. Then, as much of the tumor that can be removed without harming healthy brain tissue is taken out. Next, the skull part is replaced, and the incision is closed [32]. An endoscopic transnasal transsphenoidal approach involves removing the tumor through the nasal pathway [31]. The model suggests craniotomy only if the tumor is greater than or equal to 10mm in diameter size. Otherwise, it offers an endoscopic transnasal transsphenoidal approach.

For diagnosis as a meningioma tumor, if the tumor is found in the ventricular chambers, neuroendoscopic surgery is recommended by the model. Otherwise, if the tumor is large (10mm or more in diameter size), a craniotomy is suggested. If the tumor is small, then an endoscopic endonasal approach is recommended. Intraventricular infusion involves delivering antibodies into the brain to provide immunization against amyloid-beta peptides in the amyloid plaques that are found in AD patients [33]. Gene therapy consists in injecting brain-derived neurotrophic factors (BDNF) into the CNS to increase neuronal growth to reverse AD progression [34]. Cerebrospinal fluid shunting involves draining CSF from ventricles of the brain. This causes fresh CSF to form, which are free from tau proteins (a major cause of AD). CSF shunting is seen to decrease tau proteins by 60% in AD patient brains [35].

### 2.7 Radiation Treatment Procedures

Patients classified as having AD have been recorded to see vast improvements when exposed to low doses of ionizing radiation [42]. Advances in cognitive and motor functions were observed on AD patients exposed to the amounts of radiation [42]. Thus, NeuroXNet proposes low doses of ionizing radiation for patients who seek a non-invasive approach.

NeuroXNet recommends patients with meningioma tumors that have tumors near crucial brain structures IMRT. Otherwise, for large tumors, fractionated stereotactic radiotherapy is used. If the meningioma cannot be removed through the previous methods, then proton beam radiation is recommended to be used [44].

For patients with glioma tumors, there are two main types of radiation therapy that NeuroXNet recommends: IMRT and Image-Guided Radiation Therapy (IGRT). IGRT uses imaging software during treatment to adjust positions as needed. IMRT uses software to target radiation on the tumor and eliminate it directly. For tumors that are glioblastomic in nature, radiation therapy can be combined with the intake of temozolomide. Glioblastoma can also be treated by proton therapy [45]. NeuroXNet generates a treatment plan based on if the tumor is glioblastoma and if the patient’s tumor would be removed better through real-time imaging or not.

### 2.8 Drug Treatment Procedures

For AD patients, one of the prime medical drugs used for treatment is Aducanumab which attacks beta-amyloid proteins to reduce amyloid plaque sizes, improving cognitive function and other AD-related symptoms [38]. Other drugs that may be used include cholinesterase inhibitors like Donepezil or Rivastigmine, which improves memory and thinking functions in the brain; glutamate regulators like memantine which will enhance overall cognitive functions; and orexin receptor antagonists with suvorexant to improve behavioral symptoms [38].

The most effective drug treatment plan for patients diagnosed with PD is using levodopa with carbidopa. Levodopa is a CNS agent converted to dopamine in the brain, helping ease PD symptoms as a lack of dopamine causes PD. Carbidopa is a decarboxylase inhibitor used with levodopa and stops levodopa from being converted into dopamine before it crosses the blood-brain barrier [36]. However, this combination of drugs does not work for some patients and causes dyskinesia. This is seen to be because of DNA methylation that changes the structure of the striatal methylome or causes methyl groups to be lost, causing Levodopa-induced dyskinesia [37].

The best option for meningioma tumors is to get surgical treatment to remove their tumors. However, drug treatment can also be used with limited efficiency for those unable to get surgical treatment. The most common drugs used to treat meningioma tumors include antibody proteins like Interferon, Bevacizumab, and Hydroxyurea which prevent the growth of cancerous cells and decrease blood flow to the tumor by providing support to the immune system [39].

One of the most effective drug treatments for glioma tumors is Temozolomide (TMZ), which reduces tumor size by fastening the DNA of the cancerous cells and preventing cell division. TMZ is more effective than other drugs because it can pass through the blood-brain barrier and directly attack the cancerous tumor inside [23]. However, for patients with the O6-methylguanine methyltransferase (MGMT) protein over-expressed in glioma cells, TMZ could result in glioma cells becoming resistant and not being treated through medicine alone. This occurs because MGMT encodes a DNA repair enzyme which can cause the glioma cells to resist treatment with TMZ. TMZ treats both glioblastoma multiforme and anaplastic astrocytoma and is widely used with inhibitors of MGMT enzyme to counter any resistance that the cancerous cells build to the drug [24]. One of the most effective combinations of medications includes TMZ combined with MGMT-modified γδ T cells, which effectively prevents tumor growth and immunizes tumors through NKG2D ligands [25].

## III. Results and Analysis

### 3.1 Results for Identification of Blood Biomarkers

Five hub genes were found for PD patients with a degree of 7. These genes were CENPF, DLGAP5, KLAA0101, TOP2A, and BUB1B. CENPF or Centromere Protein F is related to the centromere-kinetochore complex and is responsible for chromosome separation during mitosis. The gene is also differentially expressed in cancer patients. DLG Associated Protein 5 codes for protein and is also found in cancerous patients. DNA Topoisomerase II Alpha (TOP2A) is crucial in the transcription process, helping create enzymes for chromosome segregation and condensation. This gene also counters drug resistance in patients with ataxia-telangiectasia [55].

For the biological function of neuron-glial cell signaling, a fold enrichment value o 58.29 was observed in 3 over-expressed PD genes. In molecular functions, the alpha1-adrenergic receptor activity process had three genes with a fold enrichment of 97.15. This shows that the over-expressed genes are essential biomarkers of PD in patients, and the extremely low false discovery rate supports that this observance is not by random chance.

For AD blood biomarkers, nine hub genes were over-expressed in the patients with the condition. RPL24 had a degree of 18, and the other eight genes each had a degree of 8, showing that these genes interacted closely and were signals of AD in patients. Ribosomal Protein L24 (RPL24) codes for protein synthesis and is part of the ribosomal proteins family of L24E. The RPL series and RPS series of genes which are the hub genes, are associated with certain cancers. The model shows that the ubiquitin ligase inhibitor activity and ubiquitin-protein transferase inhibitor activity have extremely large fold enrichments for molecular processes. These hub genes are efficient signals for AD diagnosis and can be studied for possible treatment of the disease.

For MCI patients, three hub genes were found, namely: FYN, SNRNP70, and CHD4. FYN is responsible for cell growth control in the tyrosine kinase protein family. SNRNP70 is responsible for many types of diseases involving body tissue. These hub genes are essential in classifying MCI through blood samples and can serve as a way for neurologists to differentiate between MCI and AD patients because of the different hub genes for each disease.

Five hub genes were found for patients with glioma tumors, and two biological processes of calcium ion export and regulation of postsynaptic cytosolic calcium ion concentration had high enrichment values of 38.5 and 35, respectively. PRKCA is associated with other types of cancer.

For patients with pituitary tumors, seven hub genes were found by the model. MPDZ proteins are responsible for HTR2C genes clot in the cell. Together, these hub genes can be used as non-invasive approaches for helping identify pituitary tumors in patients.

One of the most significant results of the NeuroXNet model was the biomarkers it found for meningioma tumors. The model found 37 hub genes that were overly expressed in patients with meningioma tumors, and all had degrees greater than 64. The model shows that the biological process of DNA replication preinitiation complex assembly has a high enrichment value of 78.31 among the associated hub genes. The cellular function of the cyclin B1-CDK1 complex also has a high enrichment value of 78.31. Some of the hub genes with the highest interactions included CDK1, CCNA2, AURKA, and BUB1 (also associated with PD patients). Cyclin-Dependent Kinase 1 (CDK1) has been associated with breast cancer and is involved in M-phase promoting factors. Cyclin A2 (CCNA2) is also in the same family of genes and is responsible for protein transition. The gene is also found in patients with other types of cancer.

### 3.2 NeuroXNet Classification Results

For my sequential model, NeuroXNet, the model starts with a convolutional layer with 32 filters, a stride of 2, the same padding, and a kernel size of 3. The layer takes in the input mages using an activation ReLU function. Next, the images are passed into a max-pooling layer with a pool size of 2 and a stride of 2. Then, the images pass through another convolutional layer with filter size 128. Next, the images pass into another max-pooling layer followed by a convolutional layer with filter size 128. Next, the images are passed into a batch normalization layer with a momentum of 0.8. Finally, the images are flattened through a flatten layer and are passed into the dense layer, which uses the softmax function to classify and diagnose the MRI images.

NeuroXNet was created using Keras with a Tensorflow backend and had eight layers. The model has 229,895 total parameters, which is relatively less than previously used models for disease diagnosis. The model architecture is shown below:

The model had an input of 6300 images from the Training and Validation folders, which is used to train itself. The model was run for 30 epochs and had a training accuracy of 99.79% with a loss of 0.0067 and a validation accuracy of 100% with a 0.0010 loss at the end of the 30 epochs. The training and validation accuracy and loss graphs over the epochs are shown below:

Then, the model ran the images on the test set comprising 700 images evenly split into seven classes. The confusion matrix for the testing data is shown below:

The confusion matrix shows the number of images in each class and their respective predicted values compared to the actual values from the test set. The confusion matrix also helps visualize the model’s performance on the test set through the heatmap colors. The diagonal values of 92, 97, 92, 97, 96, 95, and 94 represent the number of images that NeuroXNet correctly classified in each of the seven classes of AD, MCI, PD, Glioma, Meningioma, Pituitary, and Normal respectively. Out of the 700 images in the test set, NeuroXNet correctly classified 663 pictures, and the rest of the 37 were incorrectly classified. The few incorrect classifications could be caused by the model perceiving features of certain patient MRIs to be similar to multiple diseases making the model rely on a close probability value from the softmax function to diagnose the particular incorrect disease over the correct class. Otherwise, the model performed well, achieving an accuracy of 94.71% for multiclass diagnosis.

Other than the confusion matrix, another way to help see the model’s overall performance on the test set is the classification report which gives the precision, recall, f1-score, and support for each of the classes with macro averaging as well as the weighted averaging. The classification report for the model is shown below:

Below are the formulas used in the Classification Report:

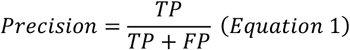

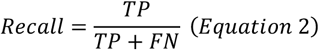

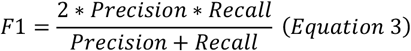

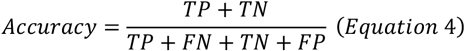

The precision, recall, and f1-score values for the model are 92%, 97%, 92%, 97%, 96%, 95%, and 94% for AD, MCI, PD, Glioma, Meningioma, Pituitary, and Normal respectively. The accuracy, macro average, and weighted average of all the different methods of calculating performance are 95%, with a support of 700.

In addition, the Cohen’s Kappa score for the model was calculated to be 0.9383 with the equation for the score shown below (p_0_ represents relating observed agreement and p_e_ represents the probability of chance agreement):

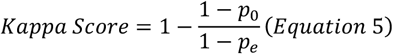

The Matthews correlation coefficient was calculated to be 0.9383 with the equation for the coefficient calculation shown below:

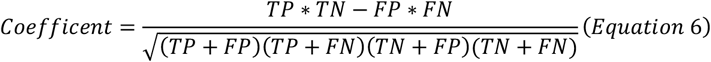

### 3.3 NeuroXNet Treatment Analysis

After NeuroXNet uses its CNN model to classify neurological diseases correctly, it uses non-imaging data inputted by the radiologist or primary physician to generate treatment recommendations. The algorithmic process that NeuroXNet uses in deciding which treatment to recommend was detailed in section 2. NeuroXNet had a 98% accuracy in predicting treatments plans with the highest survival rates.

In the final analysis, it was observed that the images of the training, validation, and testing sets all attained high accuracies and low loss values, signifying that the NeuroXNet has excellent potential in the diagnosis and treatment of neurological disorders.

## IV. Conclusion

This paper helped describe the layers and characteristics of the NeuroXNet model, which achieved a test accuracy of 94.71%. This model is the first CNN model which approaches the diagnosis of neurodegenerative diseases, primarily Alzheimer’s disease, Parkinson’s disease, and Mild Cognitive Impairment and brain tumors (glioma, meningioma, and pituitary) through a novel deep learning architecture (NeuroXNet). The model helps find new blood biomarkers for the six diseases. Furthermore, NeuroXNet is the first to integrate a treatment component into the model and uses genomic data with the MRI images to diagnose patients. Through this model, doctors and radiologists can diagnose neurological diseases at an earlier stage and use the diagnosis in treating patients with the proper medications and treatment procedures, helping prevent the disease from progressing onto a deadlier stage that could affect the patient’s health drastically. Consequently, this model has great potential to be used clinically and improve the lives of numerous patients. Through this paper, a new model is proposed and seen to attain a high accuracy that has many practical applications to radiology, neuroscience, and medicine, helping make a breakthrough in the diagnosis and treatment of neurological diseases using computational and biological approaches.

## Data Availability

All data produced in the present study are available upon reasonable request to the authors.

http://adni.loni.usc.edu/

https://www.ppmi-info.org/

https://www.kaggle.com/sartajbhuvaji/brain-tumor-classification-mri/metadata

https://www.ncbi.nlm.nih.gov/geo/query/acc.cgi?acc=GSE74385

https://www.ncbi.nlm.nih.gov/geo/query/acc.cgi?acc=GSE31095

https://www.ncbi.nlm.nih.gov/geo/query/acc.cgi?acc=GSE4488

https://www.ncbi.nlm.nih.gov/geo/query/acc.cgi?acc=GSE63063

https://www.ncbi.nlm.nih.gov/geo/query/acc.cgi?acc=GSE6613

https://www.ncbi.nlm.nih.gov/geo/query/acc.cgi?acc=GSE4226

## Notes

### Competing Interest Statement

The authors have declared no competing interest.

### Funding Statement

This study did not receive any funding.

### Author Declarations

This study involves only openly available human data, which can be obtained from: ADNI (http://adni.loni.usc.edu/), PPMI (https://www.ppmi-info.org/), Kaggle (https://www.kaggle.com/sartajbhuvaji/brain-tumor-classification-mri/metadata), and Gene Expression Omnibus (https://www.ncbi.nlm.nih.gov/geo/query/acc.cgi?acc=GSE74385, https://www.ncbi.nlm.nih.gov/geo/query/acc.cgi?acc=GSE31095, https://www.ncbi.nlm.nih.gov/geo/query/acc.cgi?acc=GSE4488, https://www.ncbi.nlm.nih.gov/geo/query/acc.cgi?acc=GSE63063, https://www.ncbi.nlm.nih.gov/geo/query/acc.cgi?acc=GSE6613, https://www.ncbi.nlm.nih.gov/geo/query/acc.cgi?acc=GSE4226).

